# Associations Between Maternal Risk Factors and Intrinsic Placental and Fetal Brain Functional Properties in Congenital Heart Disease

**DOI:** 10.1101/2022.09.06.22279657

**Authors:** Vidya Rajagopalan, Vanessa Schmithorst, Alex El-Ali, William Reynolds, Vincent Lee, Julia Wallace, Jaqueline Wienberg, Jennifer Johnson, Jodie Votava-Smith, Jennifer Adibi, Ashok Panigrahy

## Abstract

**Background:** The relationship between maternal risk factors (MRFs) (particularly pre-gravid obesity, diabetes, and hypertension) and congenital heart disease (CHD) to placental and fetal brain outcomes is poorly understood. Here, we tested the hypothesis that MRF and CHD would be associated with reduced intrinsic placental and fetal brain function using a novel non-invasive technique.

**Methods:** Pregnant participants with and without MRF and fetal CHD were prospectively recruited and underwent feto-placental MRI. Using intrinsic properties of blood oxygen level dependent imaging (BOLD) we quantified spatiotemporal variance of placenta and fetal brain. MRFs and CHD were correlated with functional characteristics of the placenta and fetal brain.

**Results:** Co- morbid MRF (hypertension, diabetes, and obesity) reduced spatiotemporal functional variance of placenta and fetal brain (p < 0.05). CHD predicted reduced fetal brain temporal variance compared to controls (p<0.05). Interaction of MRF and CHD status was associated with reduced intrinsic pBOLD temporal variance (p=0.047). There were no significant interactions of MRFs and CHD status on either temporal or spatial variance of intrinsic brain BOLD.

**Conclusion:** MRF and CHD reduced functional characteristic of placenta and brain in fetuses. MRF modification and management during pregnancy may have the potential to not only provide additional risk stratification but may also improve neurodevelopmental outcomes.

## Introduction

It has been established that feto-placental development and functioning are vulnerable to maternal risk factors (MRFs) such as diabetes, obesity, and hypertension. Impaired maternal -fetal environment, linked to these MRFs, has been shown to be associated with placental vascular malformations and dysfunction [1,2] [3]. This in turn creates an unfavorable environment for healthy fetal development. Poor neurological outcomes have been reported in offspring with prenatal MRFs [4] [5,6]. But there is very scant evidence on relationship between MRFs, placental function and brain development during the critical fetal period.

In parallel, recent histopathological evidence has reported that placentas, in CHD, show a spectrum of pathological structural adaptations such as infarctions, chorangiosis, thrombosis and hypomature villi, suggesting vascular-related vulnerability and have been linked to the presence of postnatal brain injury, particularly in left ventricular outflow track obstruction heart lesion subtypes [7-11]. While these placental pathologies have been shown to correlate with newborn outcomes such as birthweight[8] and acquired postnatal brain injury, few direct associations have been found between placental abnormalities and neurodevelopmental outcomes in CHD. Russel and Gaynor [12,13] et al recently demonstrated that damaging variants in proangiogenic genes may impact placental function and are associated with impaired fetal growth in CHD pregnancies. There is also recent evidence of overall dysregulation of placental and fetal brain angiogenesis in CHD, suggesting that these fetuses may have an intrinsic placental angiogenic impairment that could contribute to impaired functional brain development [14]. There is, also, emerging evidence that CHD together with MRF is associated with impaired placental vasculature and substantially increased risk of mortality after cardiac surgery [15-17]. Taken together, these studies support the overarching hypothesis that MRFs and CHD are with intrinsic placental and brain dysfunction.

MRI studies of placenta function have reported altered placental response in CHD pregnancies when the mother is subjected to external hyperoxia stimuli. More studies using hyperoxia stimuli have noted global reductions in perfusion and T2* in CHD placentas [18,19]. However, hyperoxygenation-induced vasoconstriction has varying effects on the intrinsic vascular properties in the placenta and the fetus[20] and therefore may mask the very differences we seek to measure. Additionally, these methods are very susceptible to motion artifacts that are inevitable in fetal imaging, require higher degrees of logistical and technological expertise making them non-ideal for translation across large scale studies. Intrinsic BOLD, which has the potential of overcoming some of these limitations, may be a physiological functional surrogate for placental vascular properties and oxygenation and vascularity, but, to date, few studies have captured intrinsic BOLD signal properties of placenta and fetal brain in CHD pregnancies.

To address this knowledge gap, we quantitated, *intrinsic* placental and fetal brain BOLD signal properties (without the use of maternal hyperoxygenation) in prospectively recruited mothers-fetal dyads with variable MRF from a single center fetal cardiology center. We developed an innovative acquisition and post-processing technique by leveraging previously validated quantitative metrics to capture both temporal (temporal variance) and spatial (spatial variance) properties of intrinsic BOLD signal in placenta and fetal brain. We recruited a sample of both CHD and non-CHD patients with varying levels of MRF. Our primary purpose was to determine the effect of MRF and CHD on intrinsic BOLD signal in the placenta and fetal brain. We hypothesize that: (1) MRF will decrease spatiotemporal variance of intrinsic BOLD signal in the placental and fetal brain; (2) Spatiotemporal variance of intrinsic BOLD signal in placenta and fetal brain will be altered in CHD compared to non-CHD.

## Methods

### Subject Population

This prospective study, conducted between 2014 and 2018, was approved by the institutional review board at the University of Pittsburgh. Table 1 shows the demographic characteristics of the participants in this study. Participants in the CHD group were enrolled from one of the following three pediatric clinics: (1) High risk pregnancy center at Magee Women’s Hospital (2) Fetal cardiology clinic at Children’s Hospital of Pittsburgh and (3) an affiliated fetal cardiology clinic (Wexford, PA). *Inclusion criteria* for the CHD group were: (1) pregnant women whose fetuses were diagnosed with complex CHD based on fetal echocardiograms; (2) in their third trimester of pregnancy -- 27-42 gestational weeks (GW); and (3) English speaking (for obtaining informed consent). Participants in the <u>non-CHD group</u> were enrolled using a variety of methods including flyers, self-referral via a university-sponsored medical research participation database and referral from fetal cardiology clinic after negative echocardiogram. *Inclusion criteria* (2) and (3) are identical to the CHD group but non-CHD participants were required to have no documented history of CHD. Patient with three major maternal risk factors including obesity, diabetes and maternal risk were not excluded. Notably, some enrolled non-CHD participants underwent fetal echocardiogram, which was normal, due to increased risk for CHD (e.g. positive family history). Exclusion criteria for both groups were: (1) Focal neurological abnormality on postnatal clinical follow up; (2) Chronic seizures post-birth; (3) Severe congenital brain malformation detected on prenatal imaging; (4) Significant chromosomal abnormality/ syndrome; (5) Sepsis or other infection; (5) Significant birth trauma and/or hypoxic ischemic injury; (6) Contraindication to MRI (e.g. claustrophobia, pacemaker).

**Table 1:** Demographic and Clinical Characteristics of CHD and Non-CHDs Groups.

| Demographics and Clinical Characteristics | Non-CHD (n=114) | CHD (n=58) | p-value |
| --- | --- | --- | --- |
| GA at MRI (wks) | 32.27 (3.01) | 33.62 (3.10) | 0.0136 |
| Mean maternal age at delivery - years (SD) | 30.69 (4.57) | 29.22 (5.94) | 0.2156 |
| Sex, n (%) |  |  | 0.0668 |
| Male | 48 (42.11) | 33 (56.90) |  |
| Female | 66 (57.89) | 25 (43.10) |  |
| Race, n (%) |  |  | 0.4461 |
| White | 87 (76.32) | 44 (75.86) |  |
| Black | 18 (15.79) | 11 (18.97) |  |
| Other or Unknown | 9 (7.89) | 3 (5.17) |  |
| Cardiac Lesion, n (%) |  |  |  |
| Hypoplastic left heart syndrome | . | 14 (24.14) |  |
| Transposition of the great arteries | . | 13 (22.41) |  |
| Double outlet right ventricle | . | 9 (15.52) |  |
| Tetralogy of fallot | . | 9 (15.52) |  |
| Complex single ventricle (TA, PA) | . | 11 (18.97) |  |
| Coarctation of aorta | . | 5 (8.62) |  |
| Other (AVC, VSD, ASD) | . | 33 (56.90) |  |

### Clinical Data

The electronic medical record (EMR) was reviewed for all participants and selected demographic variables were extracted including maternal age, gestational age (GA) at the time of the MRI. CHD lesions were classified by an experienced pediatric cardiologist (JVS) after review of pre and postnatal echocardiogram data as: (a) one of 7 categories CHD lesions listed below; (b) single versus double ventricle cardiac anatomy; (c) cyanotic versus acyanotic lesions. The seven cardiac lesion categories included: Hypoplastic left heart syndrome, Transposition of the great arteries, Double outlet right ventricle, Tetralogy of Fallot, Complex single ventricle (TA, PA), Coarctation of the aorta and Other (AVC, VSD, ASD, APVR). Cyanotic lesions included CHD lesions that were characterized by postnatal mixing of pulmonary venous and systemic venous blood resulting in reduced oxygen saturation. CHD lesions such as transposition of the great arteries (TGA), single ventricles, tetralogy of Fallot (TOF), and total anomalous pulmonary venous return (TAPVR) were considered cyanotic types of CHD lesions. Coarctation or isolated shunt lesions were considered acyanotic types of CHD lesions. Distribution of cardiac lesion types among study participants are summarized in Table 1.

### Maternal Risk Factors

Maternal risk factor data (Table 2) was collected from the Magee Obstetric Medical and Infant Database (MOMI) in conjunction with prospective data collection as part of the recruitment. In 1995, Magee-Womens Hospital established the MOMI database to integrate data on maternal, fetal, and neonatal outcomes. This data is available to researchers with institutional review board approval for secondary analyses. The Medical Records Department abstracts maternal information, and infant outcome information comes from the electronic medical records. International Classification of Diseases (ICD) 9 and 10 diagnostic information is recorded on discharge and these data are coded into the MOMI database. Clinical data specifically collected for this study included the presence of pre-gravid weight/height, diabetes, and hypertension. Patients with gestational diabetes, type 1 diabetes mellitus, and type 2 diabetes mellitus were grouped into a single “maternal diabetes” group. Patients with chronic hypertension and gestational hypertension (including pre-eclampsia) were grouped together as well. These risk factors were determined to be present if this diagnosis was indicated in the MOMI database and then checked for explicitly mentioning in a prenatal obstetrical visit note. Maternal obesity was defined as a BMI (kg/m^2^) of greater than 30 based on height and weight measurements taken prior to pregnancy (within 1 year). Pregnancy and delivery characteristics of the cohort are shown in Supplemental Table 1A. In addition, placental weight and other birth characteristics of the infant were collected from both the MOMI dataset and also the Children’s Hospital medical records (Table 3).

**Table 2:** Incidence of Maternal Risk Factors in CHD and Non-CHDs Groups.

| <u>Maternal Risk Factors</u> | Non-CHD (n=114) | CHD (n=58) |
| --- | --- | --- |
| Pregnancy Complications, n (%) |  |  |
| Any Risk | 65 (57.01) | 28 (48.27) |
| Maternal Obesity | 58 (50.88) | 28 (48.27) |
| Maternal Hypertension | 27 (23.69) | 6 (10.35) |
| Maternal Diabetes | 13 (11.40) | 5 (6.90) |

**Table 3:**
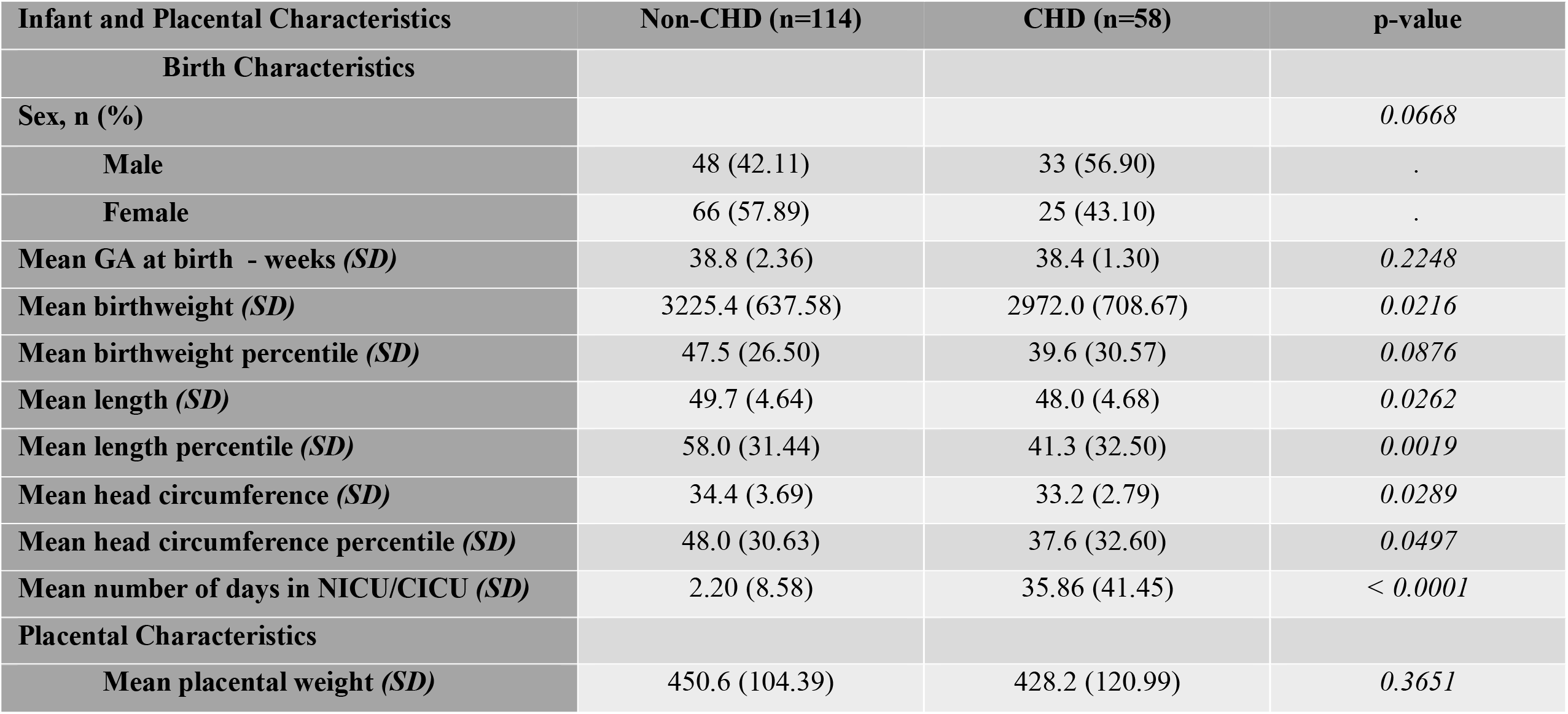
Infant and Placental Characteristics of the CHD and Non-CHD Groups.

| Infant and Placental Characteristics | Non-CHD (n=114) | CHD (n=58) | p-value |
| --- | --- | --- | --- |
| Birth Characteristics |  |  |  |
| Sex, n (%) |  |  | 0.0668 |
| Male | 48 (42.11) | 33 (56.90) | . |
| Female | 66 (57.89) | 25 (43.10) | . |
| Mean GA at birth - weeks (SD) | 38.8 (2.36) | 38.4 (1.30) | 0.2248 |
| Mean birthweight (SD) | 3225.4 (637.58) | 2972.0 (708.67) | 0.0216 |
| Mean birthweight percentile (SD) | 47.5 (26.50) | 39.6 (30.57) | 0.0876 |
| Mean length (SD) | 49.7 (4.64) | 48.0 (4.68) | 0.0262 |
| Mean length percentile (SD) | 58.0 (31.44) | 41.3 (32.50) | 0.0019 |
| Mean head circumference (SD) | 34.4 (3.69) | 33.2 (2.79) | 0.0289 |
| Mean head circumference percentile (SD) | 48.0 (30.63) | 37.6 (32.60) | 0.0497 |
| Mean number of days in NICU/CICU (SD) | 2.20 (8.58) | 35.86 (41.45) | < 0.0001 |
| Placental Characteristics |  |  |  |
| Mean placental weight (SD) | 450.6 (104.39) | 428.2 (120.99) | 0.3651 |

### Placental Pathology

Placental histopathologic and gross examinations were collected for all deliveries at our institution for which such examination was indicated clinically. Pathology examinations were performed by dedicated perinatal pathologists according to the standard clinical protocol used at our institution. Pathology reports (Supplemental Table 1B and Supplemental Table 2) were reviewed for (1) placental weight and (2) pathologic lesions including placental infarcts, vascular thrombosis, chorangiosis, and chorioamnionitis as previously described in the literature.

### MRI Image Acquisition

Maternal subjects were imaged using a 3T Skyra (Siemens, Ehrlangen Germany) with an 18-channel receive-transmit phased body array. Patients were imaged supine, if tolerated and lateral decubitus positions, depending on comfort. All patients were imaged without contrast or sedation. Placental Imaging: T2 HASTE was obtained through the uterus, for anatomic localization, with the following scan parameters: TR = 1110, TE = 78, flip angle = 142 deg. Echo-planar imaging (EPI) BOLD images were obtained through the placenta with the following scan parameters: FOV = 300mm TR = 2280, TE = 32, flip angle = 80 deg., with an in-plane resolution of 4.7×4.7 mm^2^ and slice thickness of 3 mm. 100-time frames were attempted on each mother over 3 minutes and 53 seconds. Placental BOLD images were obtained using a 3T Siemens Skyra and an 18-channel body coil. Acquisition parameters were repetition time (TR) = 4,530 ms, echo time (TE) = 32 ms, matrix = 64 × 64, field of view (FOV) = 320 × 320 mm2, slice thickness = 3 mm, 64 slices acquired covering the entire placenta. Placental BOLD sequences were repeated twice and the series with the least amount of motion observed was used for analysis.

#### Fetal Brain Imaging

Axial T2 HASTE was obtained through the fetal brain, for anatomic localization, with the following scan parameters: TR = 1110, TE = 78, flip angle = 142 deg. Echo-planar imaging (EPI) BOLD images were obtained through the fetal brain with the following scan parameters: FOV = 320mm TR = 3280, TE = 32, flip angle = 80 deg., with an in-plane resolution of 5×5 mm^2^ and slice thickness of 3 mm. 100-time frames were attempted on each mother over 3 minutes and 53 seconds.

### Image Analysis

#### Image Pre-processing

Placental imaging requires extensive pre-processing before analysis because of the unique artifacts caused by spontaneous feto-maternal motion. To ensure fidelity of data across the dataset, each image was reviewed by senior radiology resident (AE) and research associates (JW,WR) with final image review by an attending pediatric radiologist with 20 years of fetal imaging experience (AP). The first step of image analysis involved manual review of each study for imaging artifacts. Imaging artifacts primarily included (1) excessive motion of the fetus, uterus and/or mother and (2) standing wave (“dielectric artifact”).

Studies were excluded when there was excessive artifact such that post-processing techniques were likely to fail. To mitigate standing wave artifact, the placement of a saline filled pad on the anterior abdominal wall was attempted in an initial subset of patients. Ultimately, saline bag placement did not significantly affect our rate of standing wave artifact, thus it was discontinued after the first 21 patients. Next, in each image, placental region was manually extracted in the first frame of the BOLD data using Medical Image Processing, Analysis, and Visualization (MIPAV version 7.1.1; National Institutes of Health). This region was then propagated to subsequent frames to create a volume of interest (VOI) and manually assessed for fidelity. For patients in whom the entire placenta was not covered during imaging, a VOI was placed on 5 consecutive slices of the subjectively largest portion of the placenta in the axial imaging plane.

#### Computation of Temporal Variance

A custom IDL (interactive date language) program was used to calculate placental BOLD MRI temporal variance for each patient. Temporal variance was calculated by taking the average signal intensity over the placental volume for each time point and computing the standard deviation over the entire acquisition time period (Figure 1a). The values for each time point were normalized to the unity mean (mean of 1) to non-CHD for effects such as receiver gain, since MRI values are fundamentally arbitrary up to a scaling constant. Temporal variation in pBOLD is due to cyclical variations in inflow of maternal blood. During part of the cycle when maternal inflow is high, more fully oxygenated blood enters the placenta causing oxygen to be higher and lower when inflow is lower. Temporal variance is also moderated by fetal hemodynamics where oxygenated blood is flowing away from the placenta while deoxygenated blood is flowing into it lessening the effect. Thus, greater intrinsic pBOLD temporal variance is expected when oxygenation demand should be the same (fetuses of a certain GA) but vasculature is different such that inflow to the fetus is less (like in a fetus with CHD). See Supplemental Methods for further details.

**Figure 1:**
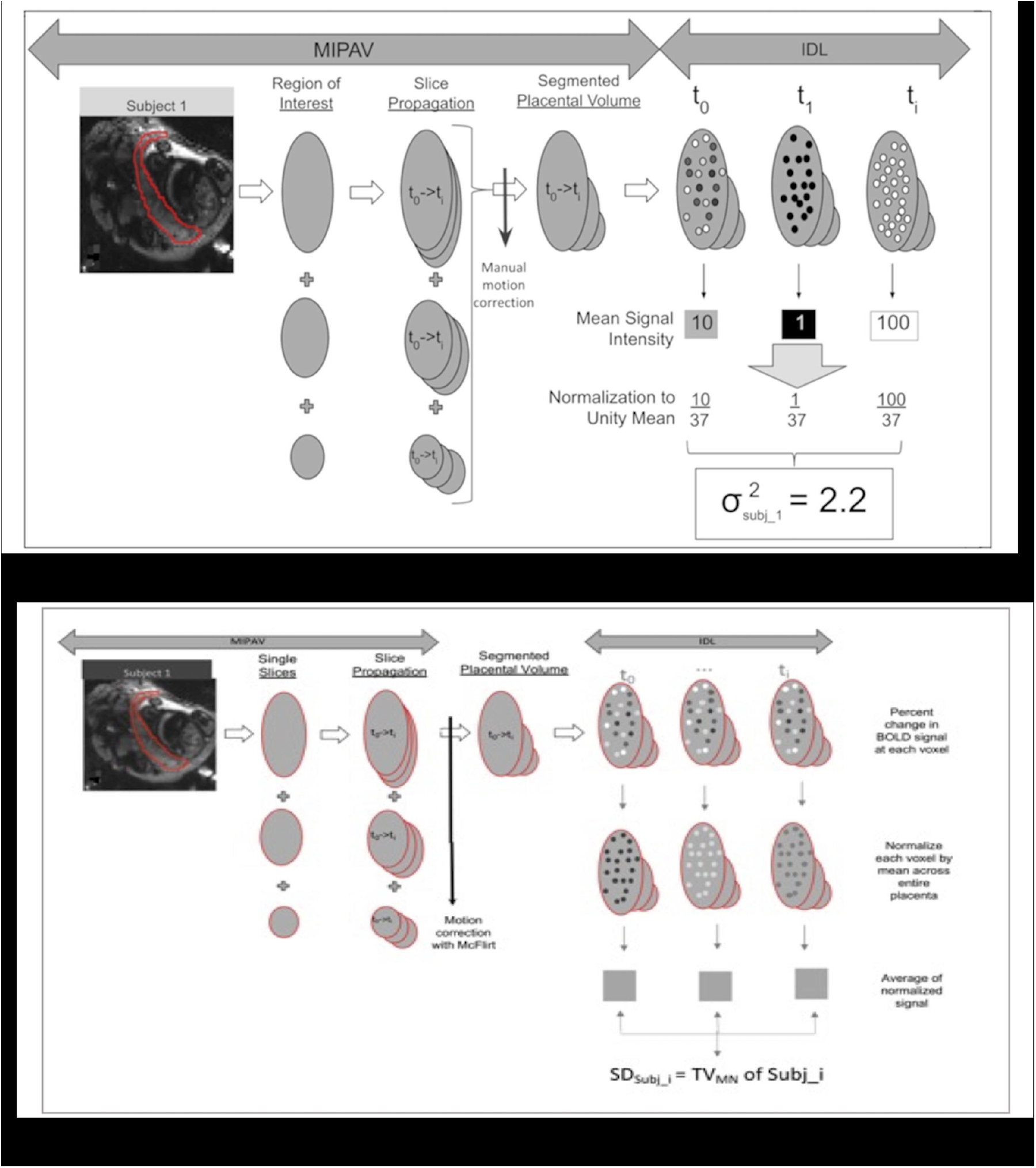
Pipeline for calculation of intrinsic spatial-temporal variance of placental and fetal brain BOLD signal: Schematics showing the computation of: (A) temporal variance (TV) and; (B) spatial variance (SV) of intrinsic BOLD signal in the whole placental structure. A similar method was used to compute these metrics in the whole fetal brain structure.

#### Computation of Spatial Variance

Following extraction of VOIs, BOLD data, were motion-corrected using an inter-slice, rigid body registration with normalized cross-correlation cost function (implemented in Python and FSL). Then, motion corrected BOLD data were corrected for noise using a combination of filtering and controlling for signal in amniotic fluid using FSL. In order to standardize the BOLD signal across all subjects, we computed the per-voxel percent change time-series[21] within the placenta. A global signal was computed by calculating the mean of percent change time-series across all voxels. The standard deviation of the global signal, defined as the mean Percentage of Amplitude Fluctuation (mPerAF[22]), corresponds to spatial variance in spontaneous pBOLD fluctuations across the placental cotelydons (Figure 1b). It is known that placental blood volume flow increased with GA to meet the increasing demands of a growing fetus[23]. In healthy placentas, spatial variance will increase with increasing GA as placental blood volume flow increases across the multiple cotelydons (See Appendix for further details). Since mPerAF is scale-independent, the spatial variance can be used to directly compared across various placentas.

#### Statistical Analysis

Student’s t-test was used to compare differences in sample means between clinical/demographic factors and placental pathology between the CHD and non-CHD cohorts. **Temporal variance:** Statistical analysis was performed using routines written in the IDL programming language. Difference in placental and brain temporal variance between CHD and non-CHDs (adjusting for gestational age) was determined by histogram analysis and calculation of the K-S test (distribution between groups) and F-test for difference in histogram measures including variance, kurtosis and skewness.. Correlation of MRFs and temporal variance of placenta and fetal brain were computed using regressions with temporal variance as the dependent variable and using a wild bootstrap (5,000 repetitions) suitable for heteroskedastic regression.[24] Spearman’s Rank-Order correlation coefficient was used to evaluate correlations between pBOLD temporal variance and brain BOLD temporal variance. Relationship between temporal variance of placenta and the fetal brain was modeled using regression analysis. **Spatial variance:** Adjusted GA: We first performed robust, multiple linear regression with spatial variance as dependent variable and GA and binary CHD status as independent variables. Maternal risk status (defined as presence of at least one of the measured maternal risks) was, then, included as an interaction variable in the previous model. Significance of relationship in the regression model were tested using the non-parametric Mann-Whitney test (p<0.05). From this point forward, any reference to significance implies statistical significance. Relative importance of individual risk factors in explaining the variance within the regression model (while controlling for GA) was computed using multivariate dominance analysis [25]. This analytic approach determines the relative proportion in which MRFs, alone or in combination, account for variance in model.

## Results

### Recruitment and Patient Characteristics

This is a prospective study of 166 mothers were recruited for fetal MRI scan. 140 patients underwent fetal imaging. 114 underwent both fetal brain and fetal placental BOLD MRI. 10 of these studies (5.5%) were not analyzable due to artifact including motion artifact (n=7), aliasing (n=1), and early imaging termination (n=2). 20 patients underwent fetal brain BOLD MRI only. 2 patients underwent only placental BOLD MRI. 4 subjects did not undergo any BOLD imaging. Among the 104 patients with adequate complete BOLD MRI scans, pBOLD was compared between CHD (n=30) and non-CHD (n=74) cohorts.

Sex, ethnicity, MRFs and placental pathology of the successfully imaged patients are found in Table 1. (sex, ethnicity, GA at fetal MRI, GA at delivery). The breakdown of cardiac lesions in the CHD cohort may be found in Table 1(cardiac lesion types, acyanotic, etc). No significant differences in ethnicity, MRF and placental pathology were observed. Within the CHD cohort, there was a greater proportion of male patients 68% vs 42% in non-CHDs group (p=0.05). GA at MRI was slightly later in the CHD group compared to the non-CHD group, 33.65 +/- 3.507 weeks vs 32.12 +/- 2.947 weeks (p=0.008). Among the non-CHD group, 26.4% of the fetuses had a family history of CHD.

### Influence of Maternal Risk Factors

#### Placenta

MRFs was associated with reduced pBOLD intrinsic spatiotemporal variance in the combined cohort (CHD and non-CHD) (Table 4). The presence of MRF, including maternal hypertension and diabetes (p=0.046, p=0.012), was associated with reduced temporal variance in the combined cohort (Table 4A). The presence of maternal risk factors, including maternal hypertension and obesity (p=0.046, p=0.0173), was associated with reduced spatial variance in the combined cohort (Table 4B).

**Table 4A:** Association between Maternal Risk Factors (MRF) and Temporal Variance in CHD and Non-CHD Group.

|  |  | Main Effect* FDR corrected |  | CHD Interaction |  |
| --- | --- | --- | --- | --- | --- |
|  | Variable | Regression Parameter | One-sided p | Regression Parameter | One-sided p |
| **Placenta** | ANY MATERNAL RISK FACTOR PRESENT | -59.05 | 0.276 | -430.58 | 0.047* |
|  | MATERNAL DIABETES MELLITUS | -90.1 | 0.012* | -70.43 | 0.258 |
|  | MATERNAL HYPERTENSION | -36.44 | 0.046* | -42.09 | 0.211 |
|  | MATERNAL OBESITY | -5.26 | 0.172 | -5.47 | 0.359 |
| **Brain** | ANY MATERNAL RISK FACTOR PRESENT | 44.75 | 0.226 | -21.79 | 0.398 |
|  | MATERNAL DIABETES MELLITUS | -66.19 | 0.05* | 97.72 | 0.117 |
|  | MATERNAL HYPERTENSION | -85.06 | 0.015* | 93.27 | 0.062 |
|  | MATERNAL OBESITY | 3.73 | 0.078 | -2.04 | 0.281 |

**Table 4B:** Association between Maternal Risk Factors (MRF) and Spatial Variance in CHD and Non-CHD Group.

|  | Variable | Main Effect* FDR corrected |  | CHD Interaction |  |
| --- | --- | --- | --- | --- | --- |
|  |  | Regression Parameter | One-sided p | Regression Parameter | One-sided p |
| **Placenta** | ANY MATERNAL RISK FACTOR PRESENT | -134.051 | 0.49 | -270.47 | 0.0532 |
|  | MATERNAL DIABETES MELLITUS | -217.44 | 0.442 | 257.57 | 0.44 |
|  | MATERNAL HYPERTENSION | -102.28 | 0.046* | -877.62 | 0.148 |
|  | MATERNAL OBESITY | -40.055 | 0.0173* | -46.65 | 0.92 |
| **Brain** | ANY MATERNAL RISK FACTOR PRESENT | -0.3814 | 0.0812 | 0.0081 | 0.987 |
|  | MATERNAL DIABETES MELLITUS | -0.7164 | 0.024* | -0.0422 | 0.958 |
|  | MATERNAL HYPERTENSION | -0.7920 | 0.008* | 0.3228 | 0.636 |
|  | MATERNAL OBESITY | 0.4037 | 0.297 | -0.9472 | 0.092 |

#### Fetal Brain

MRFs was associated with reduced brain BOLD intrinsic spatiotemporal variance in the combined cohort (CHD and non-CHD) (Table 4). The presence of maternal hypertension or pregravid diabetes was associated with reduced temporal variance in the combined cohort (p=0.015, p=0.005) (Table 4A). The presence of maternal risk factors, including maternal hypertension and diabetes (p=0.024, p=0.008), was associated with reduced spatial variance in the combined cohort (Table 4B).

### Differences between CHDs and Non-CHD Groups

#### Placenta

There were no significant differences in intrinsic spatiotemporal pBOLD signal variance between the CHD and non-CHDs groups, adjusting for gestational age and sex. While histogram characteristics of intrinsic pBOLD temporal variance, adjusting for gestational age, were not significantly different between CHD and non-CHDs (K-S test, p value=0.182) (Figure 2A), there was a significant interaction of CHD status on the relationship between gestational age and temporal variance (p value= 0.028) (Figure 2B). Likewise, there were no difference in intrinsic pBOLD spatial variance between the CHD and non-CHD groups, adjust for gestational age and sex. However, a significant negative interaction (p value = 0.004) of CHD status was noted on the relationship between gestational age and intrinsic pBOLD spatial variance (Figure 2).

**Figure 2:**
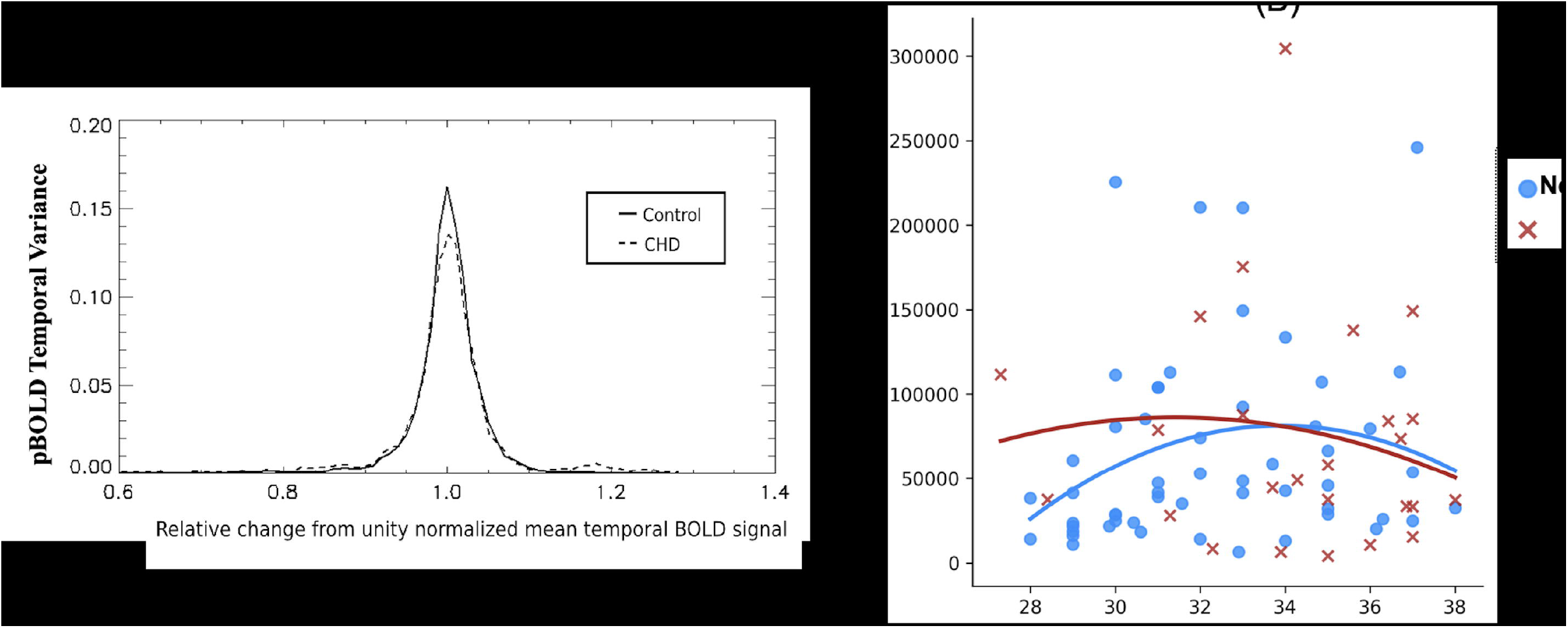
Spatiotemporal characteristics of intrinsic pBOLD in CHD and non-CHD cases: (A) Histogram characteristics of pBOLD temporal variance between CHD and non-CHD groups: while the distribution of pBOLD temporal variance between the two group is not different (K-S test, p value=0.182), the pBOLD temporal variance is reduced in the CHD group compared to non-CHDs, corrected for gestational age and ; (B) Regression plot showing significant interaction of CHD status on the relationship between gestational age and pBOLD spatial variance (p=0.004).

#### Fetal Brain

Intrinsic BOLD characteristics of the fetal brain also differed between the CHD and non-CHD groups. Temporal variance of intrinsic brain BOLD was decreased in the CHD group compared to the non-CHD group (Figure 3A). Histogram distribution analysis of non-CHD vs CHD brain BOLD curves revealed significant differences in their distribution (Figure 3A). There was no significant difference between spatial variance of brain BOLD between the two groups (Figure 3B).

**Figure 3:**
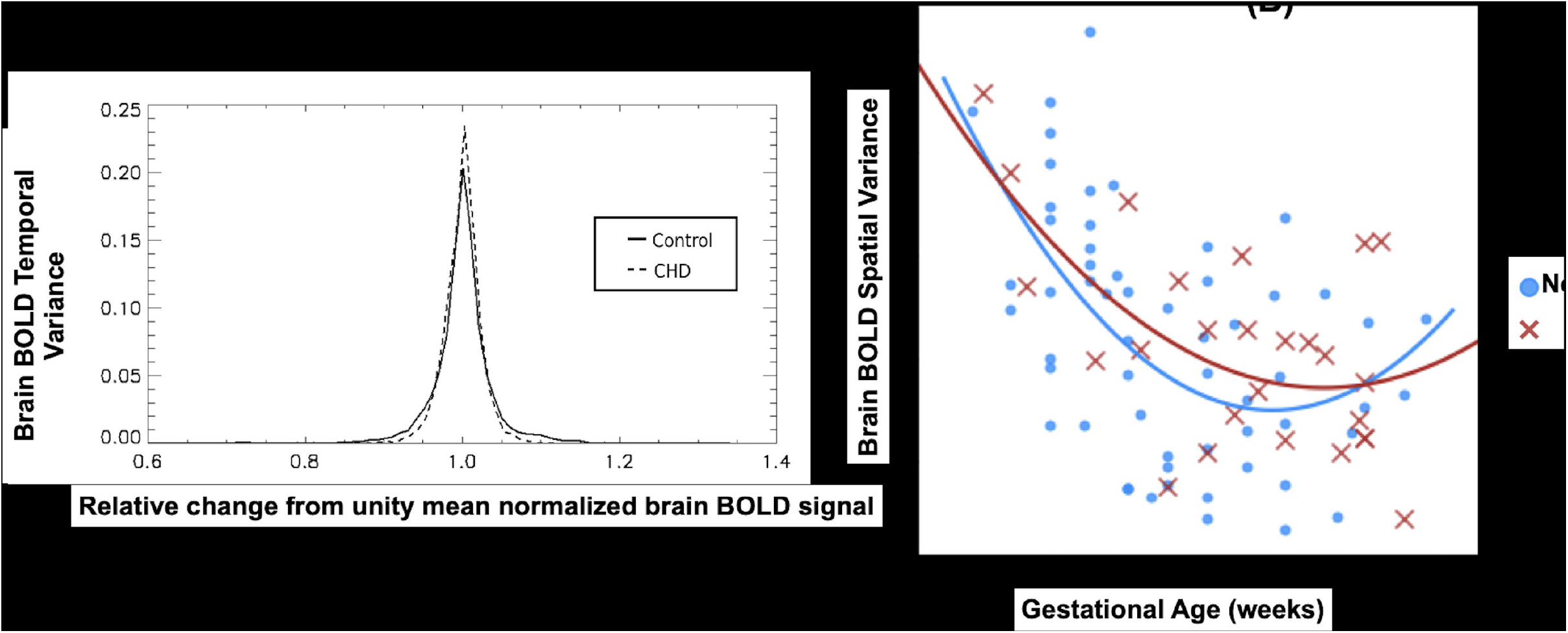
Spatiotemporal characteristics of intrinsic fetal brain BOLD in CHD and non-CHDs: (a) Distribution of brain BOLD temporal variance is significantly different between CHD and non-CHD groups, with CHD cases showing reduced temporal variance of the whole fetal brain compared to the non-CHD cases. (b) Regression plot show no significant difference in brain spatial variance with age between CHD and non-CHD groups.

### Interaction of MRFs and CHD

#### Placenta

We found a significant negative relationship between pBOLD temporal variance and the interaction of CHD and MRF (binary presence of at least one) (p=0.047) (Table 4A). There were no significant interactions of CHD and individual MRFs. There was no significant interaction of MRF and CHD on pBOLD spatial variance (Table 4B).

#### Fetal Brain

We did not find any significant interactions of MRFs and CHD status on either temporal or spatial variance of brain BOLD (Table 4).

## Discussion

In this study, our results show that MRFs is associated with reduced spatiotemporal variance of intrinsic BOLD signal in the placenta and brain in fetuses with and without CHD. Secondly, we show that fetal brain temporal variance is reduced in fetuses with CHD compared to those without. Finally, we show that interaction of MRF and CHD was associated with reduced temporal variance of intrinsic placental BOLD but not the fetal brain.

MRFs (particularly diabetes, obesity, and hypertension) are associated with increased incidence of placental impairments. Maternal diabetes has been shown to increase the incidence of placental vascularization and decreased fetal oxygenation and reduced feto-placental transfer [26,27]. Maternal obesity, which often occurs in conjunction with maternal diabetes, is associated with reduced vascularity and blood flow in the placenta[28,29]. Maternal hypertension is associated with reduced spiral artery remodeling, decreased utero-placental perfusion and placental insufficiency [30-32]. In line with these published findings, our results also show that, in the presence of these MRFs, both spatial and temporal variance is reduced in placentas regardless of CHD status.

There is a wide body of evidence showing a higher incidence of neurodevelopmental and neuropsychiatric disorders in children and adults exposed MRFs in utero [33-36]. But little is known about the direct impact of MRFs on the fetal brain. Our findings show a global reduction in spatiotemporal variance in the fetal brain in the presence of MRFs. These findings are consistent with other published work, using indirect measures of fetal brain function, demonstrating altered functional properties of fetal brains in the presence of MRFs[37,38]. These findings together suggest that the presence of MRFs is associated with altered brain organization during a critical period of development which manifests as the spectrum of neurocognitive and/or psychiatric disorders observed in offspring across their lifespan.

Significant changes to both spatial and temporal variance of intrinsic pBOLD is representative of abnormal placental vascular characteristics in CHD compared to non-CHDs. The multiple circulatory units, i.e. cotelydons, in the placenta need hemodynamic cohesion to act as an effective oxygenator meeting the increasing demands of the growing fetus. At any given GA, blood volume and flow rate are uniform in a healthy placenta. Therefore, intrinsic pBOLD signal fluctuations largely reflect changes in placental blood oxygen content. Therefore, higher pBOLD temporal variance could correspond to a less cohesive hemodynamic response among the various placental cotyledons in CHD compared to non-CHD. This could explain varying response times of CHD placentas to hyperoxygenation stimulus^42^. Differences in spatial variance, seen in Figure 2, based on CHD status, may correspond to the prevalence of regional vascular impairments in each of the cotelydons. Similar regional variations in placental perfusion in CHD were also reported by Zun [39] et al. Taking our findings together with known histopathological findings in CHD placentas [40-42], we postulate that maladaptive placental vascular development results in placental dysfunction in CHD. This alludes to the cumulative effect of CHD and increased maternal risk exacerbates feto-maternal dysfunction constituting a *“multi-hits scenario [43,44]” as* suggested by some authors.

Impaired cerebral oxygenation due to intracardiac mixing is implicated as a major contributor of deficient neurodevelopment in fetuses with CHD [45-48]. In the fetal brain, decrease in temporal variance in the CHD group is indicative of delays in the emerging functional connectivity [49,50] and is consistent with a recent study that reported delay brain function in neonates with CHD [51]. Multiple in utero factors are likely to influence fetal brain dysmaturation and long term sequalae of neurodevelopmental impairment in CHD, including deleterious gene variants, environmental factors, fetal circulatory disturbances (reduced substrate delivery including oxygen, glucose and other nutrients). Recently, placental abnormalities have been shown to play an important role in fetal brain dysmaturation in CHD patients using structural placental MRI. Andescavage et al. showed that placental volume was positively interacted with birth weight and it increased more steeply CHD-affected fetuses compared to non-CHDs [8]. Importantly, Seed et al. [52] demonstrated that decreased oxygen saturation in the umbilical venous blood, which goes from the placenta to the fetus, was associated with reduced cerebral oxygenation and impaired brain growth in fetuses with CHD compared with non-CHDs. Taken together, these studies support the concept that placental dysfunction is an important link between abnormal fetal hemodynamics and abnormal brain function in fetuses with CHD.

Our findings are consistent with the hypothesis that CHD is associated with a genetic and/or environmental background of developmental brain differences with superimposed effects of MRFs. In the fetal brain, differences in temporal characteristics between non-CHDs and CHD are reflective of alterations to the emerging functional connectivity in the brain [49,50]. Our results are consistent with what one would expect with fetal brain dysmaturation in CHD which has recently been associated with poor early neurodevelopmental outcomes [53]. However, our results further highlight the complexity of determining long-term neurodevelopmental outcomes in CHD. Further research is needed to disentangle the impact of individual MRFs on placenta and brain impairments in CHD.

Intrinsic BOLD provides a comprehensive and dynamic measure of functional characteristics in the placenta and the fetal brain. It is also more robust to motion artifacts inevitable in fetal imaging. Histopathological evidence, while useful, are subject to sampling error, do not include a measure of the total functional tissue, and occur only at a single, postnatal timepoint when functional characteristics cannot be measured. Previous examinations of functional placental and fetal brain properties have used maternal hyperoxia challenges and investigated differences in transversal relaxation time constant between the two conditions. However, placental and/or brain responses to hyperoxia challenges may not be reflective of normal function under normoxic conditions, limiting the interpretation of these type of studies.

### Limitation

Our current study did not investigate the effect of various cardiac lesion types on pBOLD characteristics due to insufficient sample size. Future multi-site studies, with larger sample sizes, can further delineate how cardiac lesion type interacts with placenta and brain BOLD characteristics in CHD. This is a cross-sectional, observational study and cannot investigate mechanisms by which MRF and CHD impair placenta and fetal brain functional characteristics.

## Conclusion

Here, we show that MRFs and CHDs differentially reduced spatiotemporal variance of intrinsic BOLD signal in the placenta and fetal brain. Additionally, we demonstrate that interaction between MRF and CHD differentially impact placental function, but not the fetal brain. Functional characteristics of the placenta and fetal brain can be measured robustly using intrinsic BOLD MRI. Targeting MRFs, via management and modification, during pregnancy provides an additional avenue for not only better risk stratification but may also improve placental and brain outcomes.

## Data Availability

All data produced in the present study are available upon reasonable request to the authors

## Abbreviations

MRI: magnetic resonance imaging
CHD: congenital heart disease
MRF: maternal risk factors
pBOLD: placental Blood Oxygen Level Dependent MRI
brain BOLD: fetal brain resting Blood Oxygen Level Dependent MRI
GA: gestational age

