## Supplemental Tables and Methods for "Associations Between Maternal Risk Factors and Intrinsic Placental and Fetal Brain Functional Properties in Congenital Heart Disease"

**Supplemental Table 1A: Pregnancy and Delivery Characteristics**

| Pregnancy and Delivery Characteristics | Non-CHD (n=114) | CHD (n=58) | t p-value |
| --- | --- | --- | --- |
| Delivery Mode, n (%) |  |  |  |
| Vaginal | 68 (59.65) | 38 (65.52) | 0.69 |
| C-Section | 41 (35.96) | 20 (34.48) |  |
| Delivery (C-Section only), n (%) |  |  |  |
| Elective | 18 (43.90) | 15 (75.0) | **0.02** |
| Labor (vaginal only), n (%) |  |  |  |
| Spontaneous | 51 (75.00) | 16 (40.11) | **0.02** |
| Induced | 17 (25.00) | 22 (57.90) |  |
| APGAR 1 minute - mean *(SD)* | 7.85 (1.83) | 8.69 (1.21) | **0.02** |
| APGAR 5 minute - mean *(SD)* | 6.98 (2.47) | 8.05 (1.91) | **0.02** |

**Supplemental Table 1B: Pregnancy and Delivery Characteristics in Relation to Placental Findings**

|  | Placental Findings | | | Placental Findings | | | Placental Findings | | |
| --- | --- | --- | --- | --- | --- | --- | --- | --- | --- |
| Pregnancy and Delivery Characteristics | **All**  **None (n=16)** | **All**  **Abnormal (n=68)** | **t**  **p-value** | **Non-CHD**  **None (n=9)** | **Non-CHD**  **Abnormal (n=38)** | **t**  **p-value** | **CHD**  **None (n=7)** | **CHD**  **Abnormal (n=30)** | **t**  **p-value** |
| Delivery Mode, n (%) |  |  |  |  |  |  |  |  |  |
| Vaginal | 11 (73.33) | 35 (51.47) | 0.21 | 5 (55.56) | 19 (50.0) | 0.49 | 6 (85.71) | 16 (53.33) | 0.08 |
| C-Section | 5 (33.33) | 33 (48.53) |  | 4 (44.44) | 19 (50.0) |  | 1 (14.29) | 14 (46.67) |  |
| Delivery (C-Section only), n (%) |  |  |  |  |  |  |  |  |  |
| Elective | 4 (80.00) | 18 (54.55) | 0.29 | 3 (75.00) | 8 (42.11) | 0.54 | 1 (100.00) | 10 (71.43) | *n/a* |
| Labor (vaginal only), n (%) |  |  |  |  |  |  |  |  |  |
| Spontaneous | 3 (27.27) | 25 (71.43) | 0.09 | 2 (40.00) | 16 (84.21) | 0.45 | 1 (16.67) | 9 (56.25) | 0.11 |
| Induced | 8 (72.73) | 10 (28.57) |  | 3 (60.00) | 3 (15.79) |  | 5 (83.33) | 7 (43.75) |  |
| APGAR 1 minute - mean *(SD)* | 8.00 (1.46) | 7.29 (2.20) | 0.13 | 8.44 (0.76) | 7.53 (2.92) | **0.03** | 7.43 (1.99) | 7.00 (2.42) | 0.63 |
| APGAR 5 minute - mean *(SD)* | 8.75 (0.45) | 8.50 (1.53) | 0.25 | 8.78 (0.35) | 8.74 (2.32) | 0.48 | 8.71 (0.49) | 8.20 (1.88) | 0.20 |

**Supplemental Table 2: Comparison of Placental Pathology between CHD and Non-CHD Groups**

| Placental Pathology | Non-CHD (n=114) | CHD (n=58) | p-value |
| --- | --- | --- | --- |
| Placental Pathology |  |  |  |
| Report available, n (%) | 47 (41.23) | 37 (63.79) | *0.0049* |
| Normal, n (%) | 9 (19.15) | 7 (18.82) | *0.9791* |
| Abnormal, n (%) | 38 (80.85) | 30 (81.08) | *.* |
| Primary placental abnormality (reported cases only) |  |  |  |
| No abnormality noted, n (%) | 9 (19.15) | 7 (18.92) |  |
| Infarct, n (%) | 10 (21.74) | 7 (18.92) | *0.7925* |
| Chorioamnionitis, n (%) | 6 (13.04) | 6 (16.22) | *0.8698* |
| Chorioangiosis, n (%) | 8 (17.39) | 5 (13.51) | *0.2727* |
| Other*, n (%) | 34 (73.91) | 26 (70.27) | *0.8373* |

*meconium macrophages, chronic villitis, hemorrhage, necrosis, thrombus, villous fibrin, accreta, subchorionitis

**Supplemental Methods:**

We assume free oxygen exchange between fetal capillaries and intervillous space (e.g., very large permeability). (If this is not the case, an adjustment can be made to the effective fetal flow rate.) We also assume completely oxygenated incoming maternal blood. We use the following variables: *V* is the total volume of the intervillous space*; H* is the hematocrit (units of heme volume / total volume)*; HV* is the heme volume of the intervillous space*; F_m_* is the maternal rate of flow (in units of heme volume / unit time) *; F_f_* is the fetal rate of flow (same units)*; D* is the deoxyhemoglobin fraction in the intervillous space (in units of volume deoxyhemoglobin / heme volume)*; D_f_* is the deoxyhemoglobin fraction in the incoming fetal blood (volume deoxyhemoglobin / heme volume), Since the deoxyhemoglobin is diffuse in the intervillous space (in contrast to BOLD fMRI where it is confined to the veins/venules) from physics the microscopic R2’ will be linearly related to the deoxyhemoglobin density:

$$R2^{'}=A*D*H$$

where A is a physical constant. Therefore, susceptibility weighted sequences will be sensitive to the parameter *D*. It is possible but not necessary to dynamically measure R2*; instead, analogous to BOLD fMRI, a single-echo gradient-echo sequence with an optimal TE may be used for optimal CNR.

From the maternal side, deoxyhemoglobin is flowing outwards (none is flowing in as the blood is assumed completely oxygenated).

$$HV\frac{dD_{m}}{dt}=-F_{m}D$$

From the fetal side, deoxyhemoglobin is also flowing outwards from the intervillous space but there is also inflowing deoxyhemoglobin from the fetal arteries:

$$HV\frac{dD_{f}}{dt}=-F_{f}D+F_{f}D_{f}$$

So the differential equation is

$$HV\frac{dD}{dt}=-F_{m}D-F_{f}D+F_{f}D_{f}$$

Which has the steady-state solution

$$D_{0}=\frac{F_{f}D_{f}}{F_{f}+F_{m0}}$$

sensitive to both fetal flow and the deoxyhemoglobin content of the arterial fetal blood.

One popular method is to employ a susceptibility-weighted sequence in combination with a maternal hyperoxygenation gas challenge. The increased maternal PaO2 is equivalent to an increase in maternal flow, producing image contrasts (between hyperoxia and normoxia) which are sensitive to fetal metabolism and placental flow on the fetal side. Indeed, single ventricle CHD shows more contrast compared to non-CHDs, since *D_f_* is higher in this population (the incoming fetal blood is less oxygenated). On the other hand, double-ventricle CHD shows less contrast compared to non-CHDs, as *F_f_* is lower. The disadvantages to this approach are the need for an invasive gas challenge and dose-response variability in the hyperoxygenation (e.g. a given % of O2 does not equate to the same PaO2 in each individual). Another approach is to take advantage of the fact that *F_m_* is time-varying as a function of the maternal circulation, and this temporal fluctuation will give rise to temporal variance in *D*. The peaks and troughs of the time evolution of *D* are given by the extrema of

$$D\left( t \right)=\frac{F_{f}D_{f}}{F_{f}+F_{m}\left( t \right)}=\frac{F_{f}D_{f}}{F_{f}+F_{m0}}*\frac{F_{f}+F_{m0}}{F_{f}+F_{m0}+\Delta F_{m}\left( t \right)}=$$

$$D_{0}*(1-\frac{\Delta F_{m}\left( t \right)}{F_{f}+F_{m0}+\Delta F_{m}\left( t \right)})$$

And thus the peak-to-peak amplitude (proportional to the temporal standard deviation) is approximately

$${D_{pp}\approx D}_{0}*\left( \frac{\Delta F_{mpp}}{F_{f}+F_{m0}} \right)=\frac{F_{f}D_{f}\Delta F_{mpp}}{{(F_{f}+F_{m0})}^{2}}$$

with similar dependence on flow and deoxygenation of fetal arterial blood (the square in the denominator is not a great difference for typical flow values of $F_{f}\approx\frac{1}{2}F_{m0}$). This approach may be done non-invasively without a gas challenge and does not have the confound of dose-response variability for the hyperoxygenation. The trade-off is that this approach is sensitive to variability in maternal systolic-diastolic placental flow differences. On the other hand, this feature may be an advantage if it is desired to investigate placental effects for maternal risk factors expected to affect maternal flow (e.g. diabetes, hypertension, etc.) Thus, we regard the two techniques as complementary.
